# COVID-19 impact on stroke admissions during France’s first epidemic peak: an exhaustive, nationwide, observational study

**DOI:** 10.1101/2021.08.03.21261438

**Authors:** Clémence Risser, Pierre Tran Ba Loc, Florence Binder-Foucard, Thibaut Fabacher, Hassina Lefevre, Claire Sauvage, Erik-André Sauleau, Valérie Wolff

## Abstract

**Background and Purpose:** The COVID-19 pandemic continues to have great impacts on the care of non-COVID-19 patients. This was especially true during the first epidemic peak in France, which coincided with the national lockdown (17 March 2020 to 10 May 2020). Patients with serious and urgent disease like stroke may have experienced a degradation of care, or may have been hesitant to seek healthcare during this period. The aim of this study was to identify, on a national level, whether a decrease in stroke admissions occurred in spring 2020, by analyzing the evolution of all stroke admissions in France from January 2019 to June 2020.

**Methods:** We conducted a nationwide cohort study using the French national database of hospital admissions (PMSI) to extract exhaustive data on all hospitalizations in France with at least one stroke diagnosis between 1 January 2019 and 30 June 2020. The primary endpoint was the difference in the slope gradients of stroke hospitalizations between pre-epidemic, epidemic peak and post-epidemic periods. Modeling was carried out using Bayesian techniques.

**Results:** Stroke hospitalizations dropped from 10 March 2020 (slope gradient: -11.70), and began to rise again from 22 March (slope gradient: 2.090) to 7 May. In total, there were 23 873 stroke admissions during the period March-April 2020, compared to 29 263 at the same period in 2019, representing a decrease of 18.42%. The percentage change was -15.63%, - 25.19%, -18.62% for ischemic strokes, transient ischemic attacks, and hemorrhagic strokes, respectively. In spatial models of French departments, the incidence of COVID-19 explained the ratio of stroke hospitalizations.

**Conclusions:** Stroke hospitalizations in France experienced a decline during the first lockdown period, which cannot be explained by a sudden change in stroke incidence. This decline is therefore likely to be a direct, or indirect, result of the COVID-19 pandemic.

## Introduction

Since the emergence of the COVID-19 pandemic, healthcare providers around the globe have been forced to mobilize their resources in order to cope in these unprecedented conditions.

The care of non-COVID patients has certainly been impacted as a result.^1–4^ The capacity of healthcare systems worldwide was rapidly overwhelmed following the beginning of the pandemic. France faced a shortage of both medical supplies and human resources during the first epidemic peak, in spring 2020, despite undertaking 55 days of national lockdown (from 17 March to 10 May 2020 included). ^5–7^ On the final day of this lockdown, the number of COVID-19 related hospital deaths recorded was 16 820.^8^ A reduction in the quality and quantity of care as a result of the extreme pressure experienced by the healthcare system may have resulted in collateral damage with regard to other patients, particularly for those with serious or urgent diseases requiring immediate treatment.^1,9,10^ In addition, many patients have been hesitant to seek heath care during this period, a phenomenon briefly described in the literature regarding the shorter SARS epidemics.^11,12^ To date, several works have alerted to an unexpected decrease in hospital admissions for non COVID-19 diseases, but very few have estimated this on a national level.^13–15^ The aim of this study was to determine whether the number of hospital admissions for stroke in France decreased during the COVID-19 epidemic peak and lockdown period, by analyzing the evolution in number of all stroke hospitalizations from January 2019 to June 2020. The secondary objectives were to observe the evolution of different types of stroke; to compare selected hospitalization characteristics between the epidemic peak period March-April 2020 and March-April 2019; to study the correlation between the incidence of hospital admission for COVID-19 and the evolution in stroke admissions between March-April 2020 and March-April 2019 by French department.

## Methods

The data that support the findings of this study are available from the corresponding author upon reasonable request.

### Ethical approval

This study adheres to French legislation regarding the reuse of anonymized data (MR-005 of Commission Nationale de l’Informatique et des Libertés), and is registered at Strasbourg University Hospital under the reference 21-2019. Relevant data files are deposited on the Health Data Hub. The Strasbourg University Hospital Ethics Committee has approved this study (reference: CE-2021-14).

### Study design and setting

Anonymized data were extracted from the French national “Information Systems Medicalisation Programme” (PMSI) database, which includes all hospitalization data transmitted by all public and private hospitals in France. Diagnoses are coded using the International Classification of Diseases, 10th Revision (ICD-10). In France, at the end of a hospital stay, all patient diagnostics and medical procedures are recorded from the patient’s medical records according to national coding rules. Medical and administrative information is transmitted monthly to the national database within one month of the patient’s discharge. A stroke diagnosis is defined by the French national agency for the management of hospital admissions data (Agence Technique de l’Information sur l’Hospitalization, ATIH) as one of the following codes: subarachnoid hemorrhage (I60.-), intracerebral hemorrhage (I61.-), other nontraumatic intracranial hemorrhage (I62.-), cerebral infarction (I63.-), stroke not specified as hemorrhage or infarction (I64.-), transient cerebral ischemic attacks and related syndromes (G45.-). We have defined three different categories of stroke for the purposes of this study: “hemorrhagic” (I60, I61, I62), “ischemic” (I63), and “transient” (G45). The database request, made retrospectively on 13 October 2020, included all hospital admissions between 1 January 2019 and 30 June 2020, with at least one stroke diagnosis, as defined by ATIH coding, during the stay. In order to prevent any selection bias which may occur by counting the same acute episode of stroke twice, only one admission per patient was counted if two admissions for the same patient were separated by one day or less. The study size was therefore defined by the relevant entries extracted from the database. The age of the patient was recorded as the age upon admission to hospital. The location of hospital stay was determined by the French department in which the patient was initially admitted.

The primary endpoint is the difference in slopes of stroke hospitalizations between the different periods (pre-epidemic, epidemic peak, post-epidemic) estimated by the model. The same model estimates the 3 knots (the number of knots being arbitrarily defined), the 4 segments and the slopes.

The secondary endpoints were the difference in the number of stroke admissions between March-April 2019 and March-April 2020: total number of strokes, number of each stroke subtype (ischemic, transient, hemorrhagic), proportion of men affected, proportion of deaths following hospitalization.

### Statistical methods

Quantitative variables are presented as the mean with standard deviation, or median with the first and third quartiles of the distributions. Qualitative variables are presented as numbers and percentages.

We considered the change in the number of stroke admissions as centered 7-days rolling means. To compare slopes between time periods, we modelled the evolution of stroke hospitalizations in four segments of lines (stability, fall, rise, return to baseline) joined by three estimated knots. Disregarding possible seasonality effects, we assumed relative stability of hospitalizations, including during the summer of 2019, before a sudden change in 2020.

Modeling was carried out using Bayesian techniques with a 95% credibility interval. Probability of superiority is rounded at both extremities to <0.0001 and >0.9999. The prior distributions are assumed to be very uninformative except for the position of each of the 3 knots, which is assumed to be uniformly distributed over the time interval between 15 February 2020 and 30 June 2020.

To identify a cause behind the change in the number of stroke admissions, a spatial correlation study was carried out by graphically representing, by French department, the incidence rates of hospital admissions for COVID-19 in March-April 2020, and the ratio between the number of stroke admissions in March-April 2020 compared to those in March-April 2019. The incidence rates of hospital admissions for COVID-19 were standardized by age and sex, using the official data estimates of French department populations by quinquennial ages and gender in 2020, published by the National Institute of Statistics and Economic Studies (INSEE)^16^; if two stays for COVID-19 for the same patient were separated by 1 day or less, only one stay was counted.

An ecological normal regression was fitted on data collected from metropolitan France between the ratio of stroke hospitalizations to standardized incidence for COVID-19, and a quadratic spatial trend established (on the centroids of each department), in order to adjust for spatial effects.

We used SAS Enterprise Guide 8.3 for data requests on the PMSI national database. All statistical analyses and graphs were carried out using R software, version 4.0.2, R Core Team (2020).

## Results

From January 1, 2019 to June 30, 2020, there were a total of 249 013 hospitalizations for stroke in France. The number of strokes over the period March-April 2019 was 29 263, compared to 23 873 during the same period in 2020. The curve representing the number of stroke admissions over this period smoothed by the 7-day average, demonstrates that with the exception of a slight decrease during the summer period of 2019, the number of strokes remains constant until a drop in March 2020 (Figure 1).

**Figure 1.**
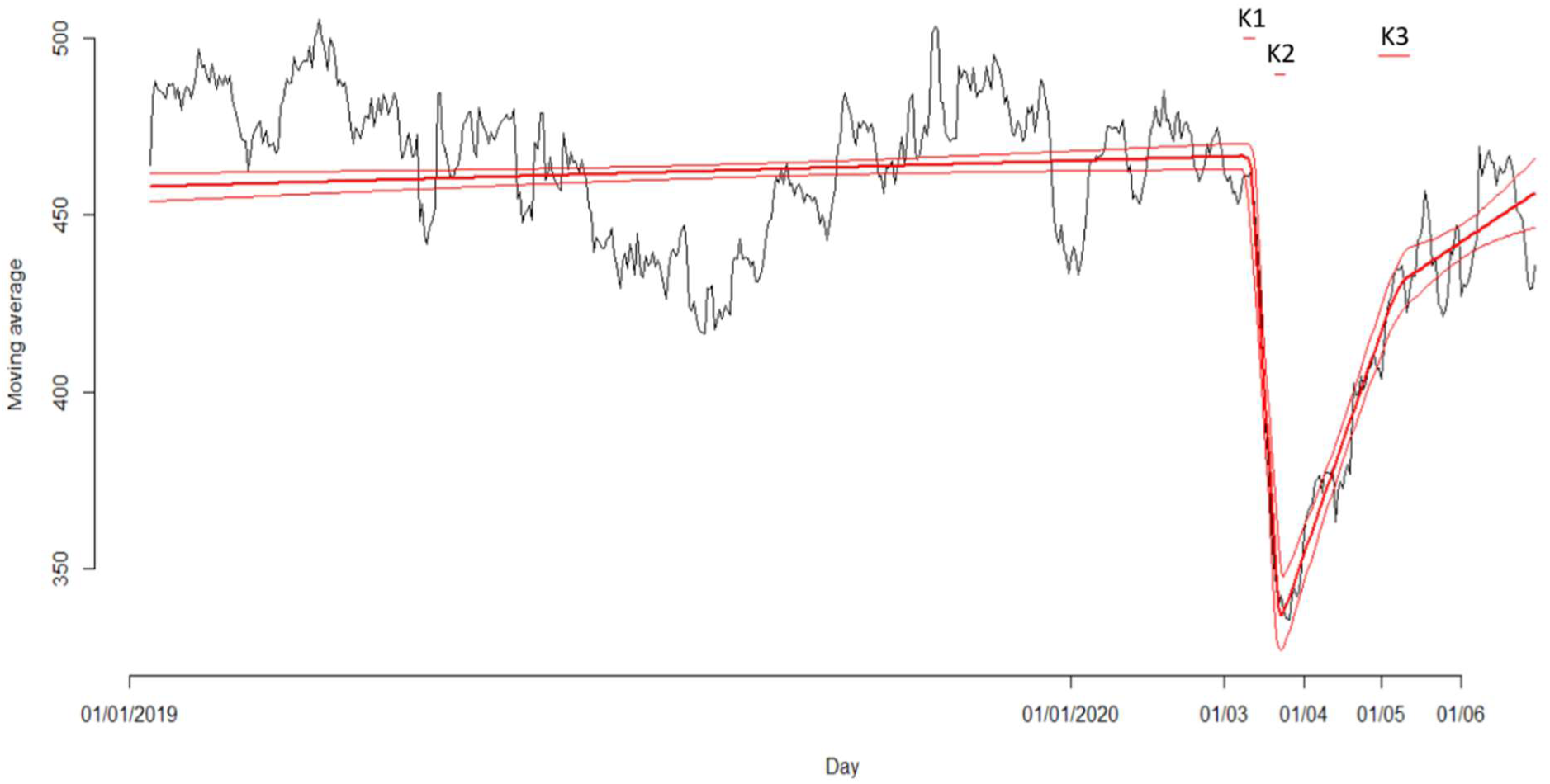
Evolution of the number of hospital admissions for stroke in France from 1 January 2019 to 30 June 2020. A sudden drop in hospital admissions for stroke can be seen in March 2020. Data is centered by 7-days rolling means, and modeling of the posterior median is represented in red with its 95% credibility interval. Horizontal short lines represent the 95% credibility interval of the three knots K1 (10 March 2020), K2 (22 March 2020) and K3 (7 May 2020).

The posterior medians for three knots were estimated on: 10 March 2020 (K1), 22 March 2020 (K2) and 7 May 2020 (K3), with small credibility intervals (K1, K2, K3 and their credibility intervals are detailed in Table 1, and credibility intervals shown as horizontal lines at the relevant position in Figure 1). Because the 7-day rolling mean number of hospital admissions is modeled, the changes in hospitalizations may have occurred in the three days either side of the given date. After 10 March 2020 (K1), the slope of the second “fall” segment was negative, at -11.70 [-16.25; -8.552]. At K3, i.e., after the “rise” period, the mean number of hospital admissions is still -37.26 per day compared to the number calculated if the slopes of all segments had remained the same as that of the first segment. After 7 May 2020 (K3), the slope of the fourth segment remains higher than that of the first segment (posterior median 0.4663), resulting in an upward slope compared to the “stability” period, albeit lower than that of the third “rise” period (2.090).

**Table 1.** Quantitative results obtained by modelling the evolution of hospital admission numbers for stroke.

|  | Posterior<br>median | 95% credible<br>interval | Probability<br>of<br>superiority |
| --- | --- | --- | --- |
| Slope segment 1 | 0.02012 | [0.004876 ; 0.03655] | 0.9953 |
| Slope segment 2 | -11.70 | [-16.25 ; -8.552] | <0.0001 |
| Slope segment 3 | 2.090 | [1.772 ; 2.470] | >0.9999 |
| Slope segment 4 | 0.4863 | [0.1883 ; 0.7784] | 0.9991 |
| Difference between slopes 4 and 1 | 0.4663 | [0.16594 ; 0.7585] | 0.9984 |
| Difference between slopes 3 and 4 | -1.611 | [-2.132 ; -1.105] | <0.0001 |
| Knot 1* | 429.9 | [427.4 ; 431.7] |  |
| Knot 2* | 441.2 | [439.4 ; 443.3] |  |
| Knot 3* | 487.8 | [479.7 ; 491.5] |  |
| Delay (in days) between knot 2 and<br>knot 1 | 11.34 | [8.253 ; 15.28] | >0.9999 |
| Delay (in days) between knot 3 and<br>knot 2 | 46.64 | [38.30 ; 49.84] | >0.9999 |
| Difference in mean number of<br>hospitalizations at knot 3 | -37.26 | [-51.09 ; -26.78] | <0.0001 |

\*K1, K2, K3: in days since January 4, 2021

In Figure 2, showing the evolution according to the type of stroke, the decrease in hospital admissions is visible for all three types of stroke during the period March-April 2020. The number of admissions in March-April 2019 was 16 776, 6978 and 5509, respectively, for ischemic, TIA and hemorrhagic stroke, compared to 14 154, 5220, and 4483 for the same period in 2020. As shown in Table 2, decrease in hospitalizations is common to all subtypes of hemorrhagic stroke, in spite of their differing physiopathologies and care.

**Figure 2.**
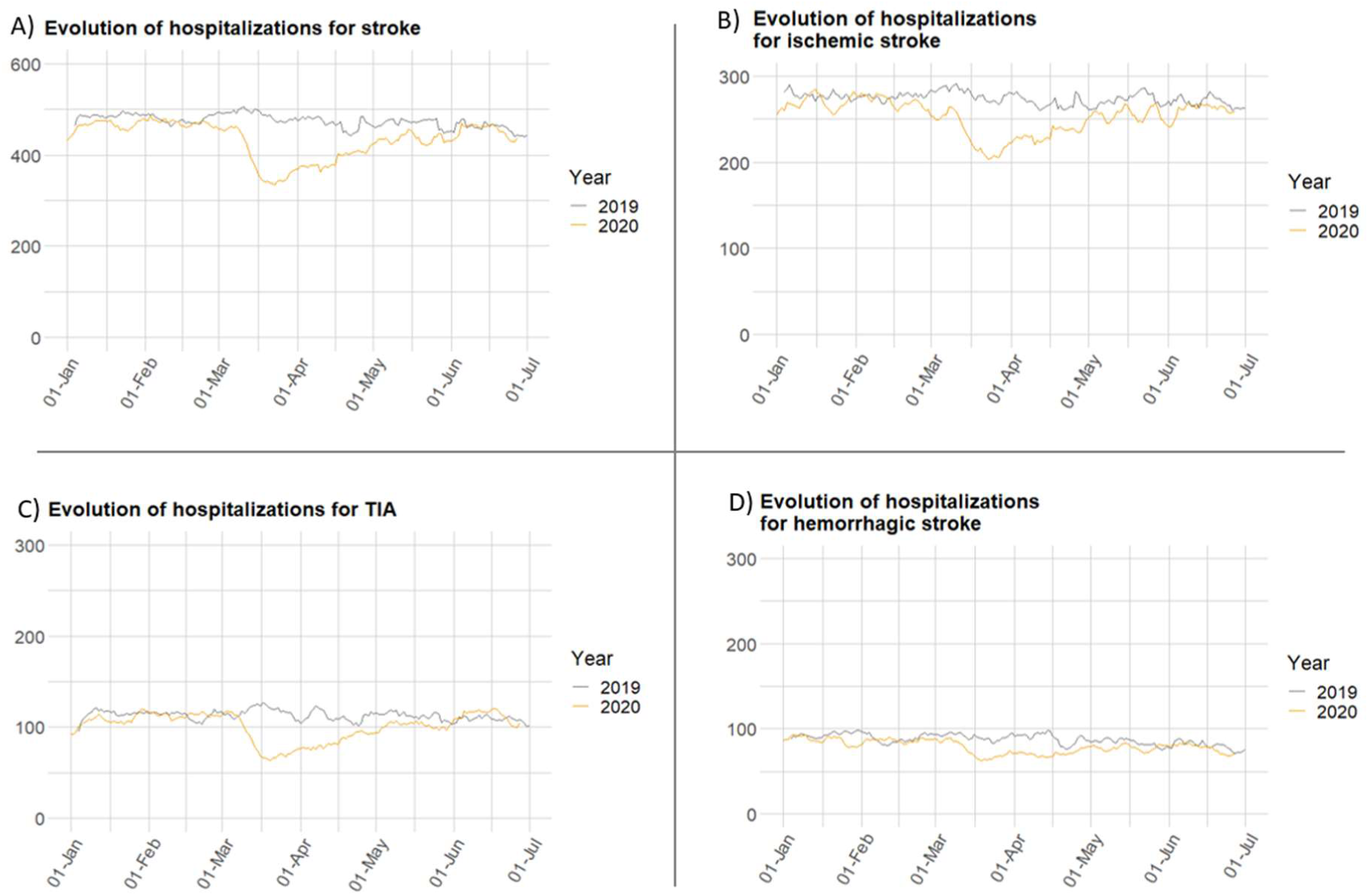
Evolution of the number of hospital admissions for different classes of stroke in France. The evolution of the number of hospital admissions centered by 7-day rolling means, from January to June is shown for (A) all types of stroke, (B) ischemic stroke, (C) TIA stroke, (D) hemorrhagic stroke. Data for the year 2020 is shown in yellow and data for the year 2019 is shown in gray.

**Table 2.** Characteristics of stroke hospital admissions.

|  | **March-April**<br>**2019** | **March-April**<br>**2020** | **Evolution**<br>**2020/2019 (%)** |
| --- | --- | --- | --- |
| **Age (years)** |  |  |  |
| Median (Q1, Q3) | 75 (64, 85) | 75 (64, 85) | 0 |
| Mean (S.D.) | 72.77 (15.52) | 72.77 (15.33) | 0 |
| **Duration of hospital stay**<br>**(days)** |  |  |  |
| Median (Q1, Q3) | 7 (3, 13) | 6 (3, 11) | -14.29 |
| Mean (S.D.) | 10.24 (13.07) | 9.090 (10.50) | -11.23 |
| **Proportion of men** |  |  |  |
| Number of men (% of total) | 15059 (51.46) | 12355 (51.75) | 0.56 |
| **Admissions ending in**<br>**death** |  |  |  |
| Number of deaths (% of<br>total) | 3032 (10.36) | 2661 (11.15) | 7.63 |
| **Number of admissions by**<br>**stroke subtype** |  |  |  |
| All | 29263 | 23873 | -18.42 |
| Ischemic | 16776 | 14154 | -15.63 |
| TIA | 6978 | 5220 | -25.19 |
| Hemorrhagic | 5509 | 4483 | -18.62 |
| Stroke not specified as<br>hemorrhage or infarction | 1050 | 773 | -26.38 |
| **Number of hemorrhagic**<br>**strokes by subtype** |  |  |  |
| Subarachnoid hemorrhage | 1148 | 942 | -17.94 |
| Intracerebral hemorrhage | 3472 | 2847 | -18.00 |
| Other non-traumatic<br>intracranial hemorrhages | 1145 | 918 | -19.83 |

S.D. = standard deviation; Q1 = lower quartile; Q3 = upper quartile

Hospital stays were shorter in March-April 2020 than in March-April 2019, whilst the proportion of men, as well as the ages of admitted stroke patients, were comparable between the two years (Table 2). The death rate was slightly higher in 2020. In March-April 2020, 2.26% of hospitalizations for stroke were associated with a COVID-19 diagnosis.

As shown in Figure 3A, the ratio of the number of stroke admissions in March-April 2020 to the number of stroke admissions in March-April 2019 was lower than 1 for 89 out of 96 departments (metropolitan France only). The departments with the greatest decrease in admissions are the region of Paris, the north east and the south east of France. The map of age and sex standardized incidence rates of COVID-19 hospital admissions by department (Figure 3B) shows a higher incidence in the north east of France and the Parisian region.

**Figure 3.**
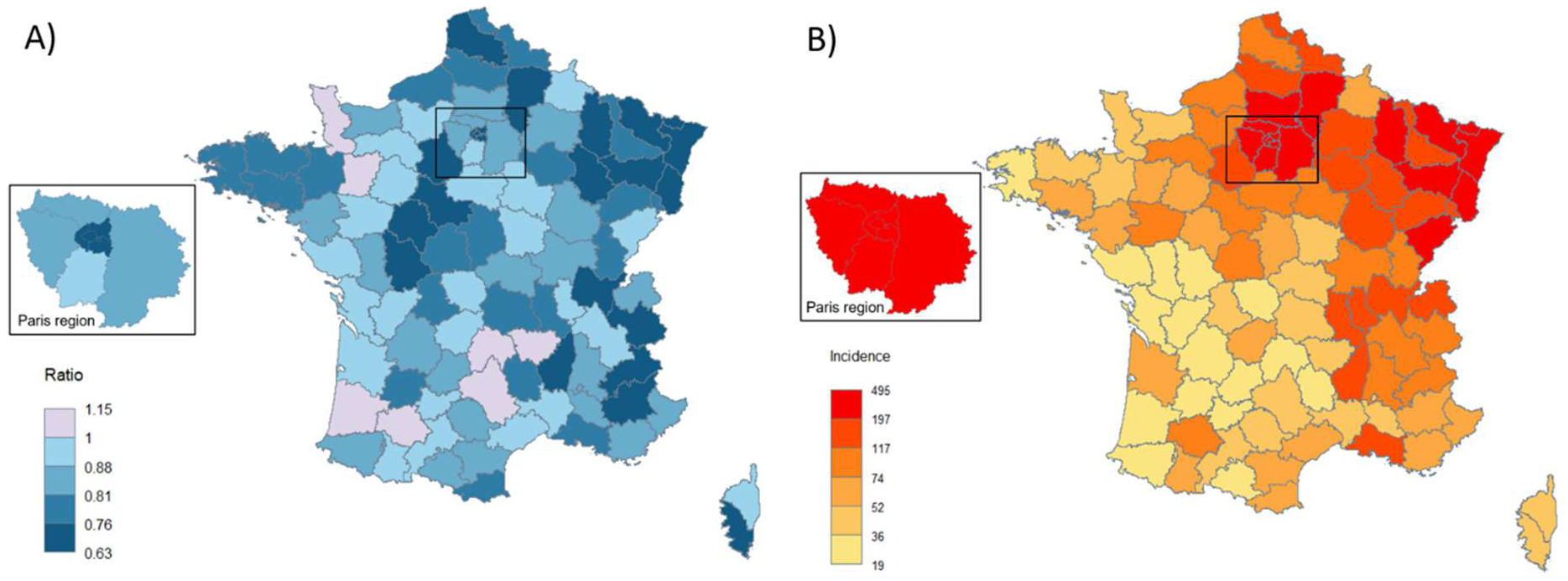
Maps of France by department. (A) The ratio of stroke hospital admissions in 2020 and those in 2019 is shown by department. Departments are colored according to the calculated ratio: pink if 1 or above; blue if lower than 1, with darkening shades of blue indicating lower ratio values. The ratios lower than 1 have been classed into quartiles. (B) The incidence of COVID-19 hospital admissions, standardized by age and sex, is shown for each department. Departments are color coded from yellow to red, with darkening shades of red representing a higher incidence of hospitalizations. Classes of the legend are not defined, automatic method is used corresponding to: nclass = 1+3.3*log10(N), where nclass is the number of class and N is the variable length.

Spatial models showed that the incidence of COVID-19 hospitalizations explains the ratio of stroke admissions (as the zero line is outside of the credible interval of the smoothed effect), whether the spatial trend is included in the model or not (Figure 4).

**Figure 4.**
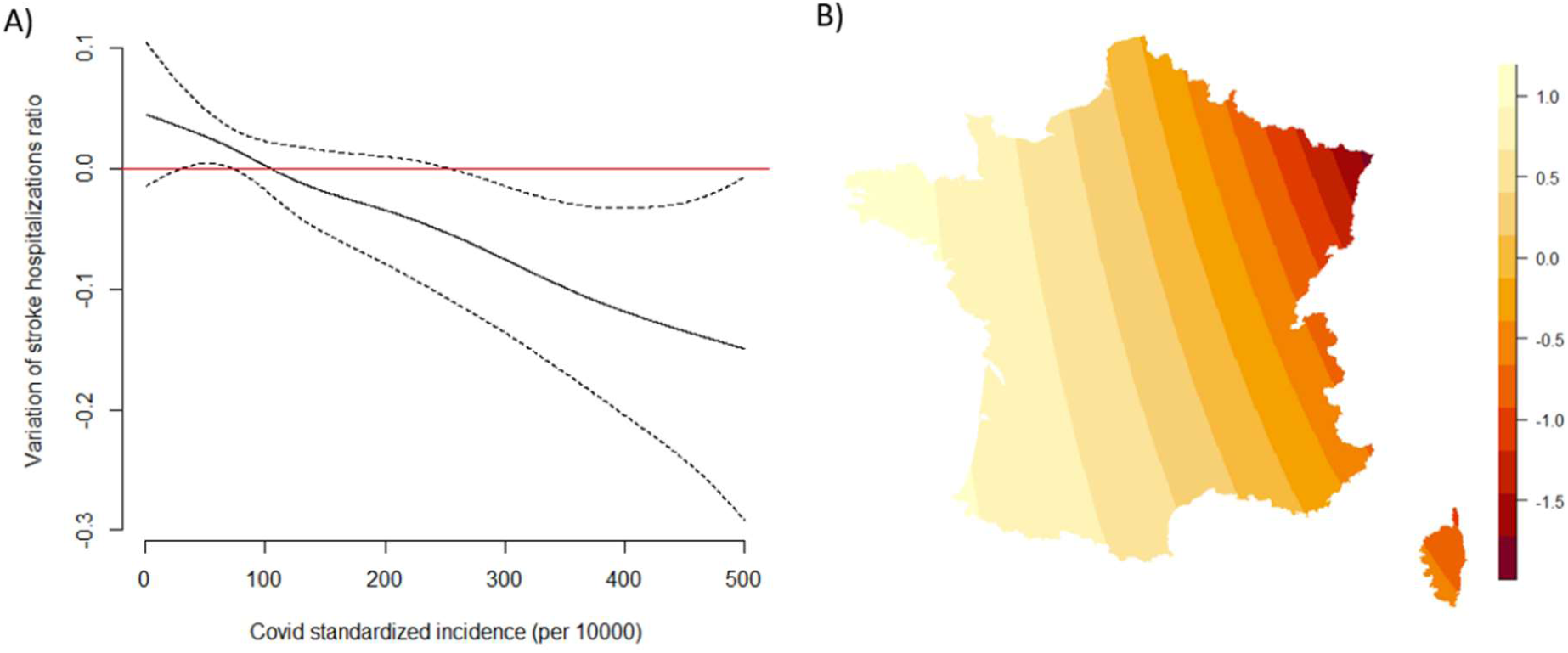
Effect of the incidence of COVID-19 hospital admissions on the stroke hospitalization ratio (March-April 2020: March-April 2019) and estimated spatial quadratic trend to adjust for a potential spatial effect. (A) shows that the reduction of the ratio correlates with an increase in standardized COVID incidence. (B) Map of estimated spatial quadratic trend in metropolitan France, shaded from light yellow (lower reduction of ratio) to dark red (stronger reduction). Because of the different spatial scales between aggregated data for French departments and the continuous quadratic trend, the unit in the map is an estimation of ratio reduction.

## Discussion

We showed that the number of patients admitted to hospital for stroke decreased significantly in France during the peak of the COVID-19 epidemic, at the time when a pandemic was declared by the WHO and lockdown measures were taken in France. This decrease is seen across all stroke subtypes. Proportions between the sexes were similar between the two periods. Differences found in the duration of hospital stay and the mortality rate are difficult to interpret because some long-term hospital stays in 2020 may have ended only after data extraction, and/or relevant data may have been uploaded late to the national database. In contrast, long-term hospital stays in 2019 are much less likely to be affected by such factors, as a longer follow-up period is present.

This sudden drop in the number of hospital admissions cannot be explained by a change in incidence due to seasonality, nor by a change of hospital facility, because our study is national. It is most likely a direct, as well as indirect, consequence of the COVID-19 pandemic. The apparently identical spatial distribution between COVID-19 and decreased stroke admission does not seem to be due to an identical spatial trend, but rather to the influence of COVID-19 on stroke admissions, because the link between the two persists even when a trend is also modelled.

This phenomenon was described in the literature for emergency department visits during the 2003 SARS outbreak in Taipei (up to 51.6% reduction in the number of daily emergency department visits compared to the number of visits prior to the outbreak).^12^ It was also described for emergency department visits in Toronto during the 2003 SARS outbreak (21% reduction over the 4-week study period). ^17^ More recently, authors described the impact of lockdown measures on emergency department visits in Lebanon, determining a 97.11% decrease per day following the “declaration of general mobilization”.^18^ Equally, in a tertiary pediatric emergency department in Cincinnati, United States, an area which experienced relatively low rates of COVID-19, it was found that the overall volume of patients in the emergency department and emergency care decreased at a daily rate of -19.4% (95% CI [-22.6%, -14.1%]) after the launch of local public health interventions against COVID-19.^19^ Few works published in 2020 have focused on the decrease in hospital admissions for stroke during the pandemic, concentrating mainly on emergency department visits, or have done so only in limited geographical areas. ^14,15,20,21^ In 2021, Richter et al showed a nationwide decrease of strokes in Germany, and a worldwide study by Nogueira et al on 457 stroke centers found an 11.5% decline in stroke admissions between the months prior to the pandemic and during the pandemic.^22,23^ Siegler et al. hypothesize that the decrease in stroke admissions is a consequence of patients presenting milder symptoms, who are therefore less likely to seek healthcare during the epidemic.^14^ This could be the result of patient apprehension regarding nosocomial infection or the feeling of being an extra burden on healthcare professionals in times of crisis. The reduction in stroke admissions could equally be the result of healthcare providers redirecting a higher proportion of patients out of hospital due to a lack of capacity, or, due to an overall under-diagnosis of stroke in these unprecedented and demanding pandemic conditions. In support of these hypotheses, our study found a greater decline in stroke admissions in several departments which experienced a higher incidence of COVID-19 hospitalization.

It seems unlikely that the epidemic context associated to national lockdown could have influenced stroke risk factors within such a short time. In the literature, the incidence of neurological symptoms in patients with COVID-19 is between 1.6% and 2.5%, depending on the study.^24–28^ A bicenter study of two New York City hospitals found that 31 patients out of a total 1916 admitted to the emergency department or hospitalized with COVID-19 had suffered an acute ischemic stroke, including 8 in which stroke was the reason for referral. In comparison, of the 1486 patients with a diagnosis of influenza, 3 had suffered an ischemic stroke. The authors found that even after adjustment for age, sex, and race, there is a higher probability of stroke with SARS-CoV-2 infection than with influenza (OR 7.6; 95% CI 2.3-25.2).^28^ The relationship between COVID-19 and stroke therefore exists, and a history of stroke results in a worse prognosis for a patient with COVID-19. The incidence of stroke in COVID-19 patients is difficult to assess, and causality difficult to establish, but it appears to be in favor of increasing rather than decreasing the incidence of stroke. This may be due to the pathophysiology of COVID-19, which involves inflammation and hypercoagulability.^26,28– 33^ Although COVID-19 may be responsible for neurological symptoms, it is also possible that it may have been a concurrent cause of death.^32^ As a result, a number of patients who may have otherwise suffered a stroke may have died from COVID-19 before the stroke occurred, or indeed, may have developed a stroke but which remained undiagnosed for a variety of factors linked to the health crisis context.

To the best of our knowledge, this is the first nationwide study in France, a country with a relatively high incidence rate of stroke (in 2014, the acute care hospitalization rate was 167.9 per 100,000 habitants, age-standardized according to the 2010 European population^34^), describing the evolution of stroke admissions during the first epidemic peak of COVID-19. Thanks to a time period of few months, we have an almost exhaustive count of stroke hospitalizations in France from January 2019 to June 2020. Using our statistical methods, we have been able to highlight the decline in national stroke patient care during the peak of the epidemic, while taking into account the evolution of stroke admissions over the entire year.

These results alert us to the need for adapting stroke management in such circumstances, and, more broadly, how to ensure appropriate care for non-COVID-19 patients during the epidemic. The impact of the epidemic on cerebrovascular diseases may be seen in the months and years to come. In contrast to other studies, we did not restrict the analyses to stroke emergency department visits, but to hospital admissions for stroke, which is even more alarming. In addition, to measure the magnitude of the impact on stroke, we decided to count admissions rather than patients, taking into account the possibility that patients may suffer multiple strokes during the study period. We also pooled transfers between hospitals during patient care to avoid a stay being counted several times.

A limitation of using the PMSI national database is that a possible delay can occur between patient discharge and coding. However, the extent of this bias is limited, as the hospital is reimbursed by the state health insurance only if diagnoses made during hospitalization are sent to the national database no more than one month after discharge. To ensure that data is as complete as possible, the analysis period was ended on 30 June 2020, i.e., leaving more than three months for patient codes to be registered before the date of extraction from the database in October 2020. Homogeneity of the diagnostic coding, which must be as close as possible to those noted during the stay, is provided by strict, national rules with regular checks carried out by the payer, thus limiting misclassification bias. A final limitation of our study is restricted knowledge of other external local events which could influence the activity of the hospital facility. Such biases are nevertheless partially controlled by the study design, as analysis is carried out at a national level, and the annual time periods compared time periods compared are proximal.

Within this study, we have not extended the analysis across a sufficient number of previous years to be able to determine whether there is a seasonal effect on stroke admissions. In previous unpublished analyses, carried out for internal purposes using data from Strasbourg University Hospital, we were able to identify a decrease in the trend of hospital admissions for stroke during the summer months of July-August, from 2015 to 2019. It is unclear whether this potential seasonal effect also concerns the period of March-April, in which case it would reduce the effect size of the difference measured during the epidemic peak.

## Summary/Conclusions

This study has determined a nationwide decrease in stroke hospital admissions at the time of the first COVID-19 wave in France. The drastic lockdown measures and unprecedented epidemic context have most likely impacted the probability of patients seeking hospital assistance in France, particularly in those regions most affected by COVID-19. In light of these findings, the care provided for stroke should be reconsidered in order to prevent stroke under-diagnosis, to improve outpatient medical care, and to facilitate health provider decision-making during the crisis. State news conferences announcing restrictions should also emphasize the importance of continuing to seek expert medical care when needed, for example, in the case of stroke. It should be highlighted to the general population that patient care for stroke remains of high quality during the crisis, and that the emergency services should always be contacted if signs of stroke appear. Equally, the public health authorities should take into account the direct, and indirect, consequences of restrictive measures on stroke admissions, and use this to inform public health decisions. It would also be informative to look at the long-term effects of the epidemic on stroke admissions.

## Non-standard abbreviations and acronyms

PMSI: Programme de Médicalisation des Systèmes d’Information (French national hospitalization database)
ATIH: Agence Technique de l’Information sur l’Hospitalisation (French national agency for the management of hospital admissions)

## Acknowledgments

CR, VW and EAS designed the study

PTBL and CR extracted the data

CR, PTBL, EAS, VW, FBF, HL, CS analyzed the data

EAS, CR, TF and PTBL performed the statistical analysis

CR wrote the manuscript

All authors reviewed, revised and approved the final report

Manuscript edition was provided by Dr. Kate Dunning (Hôpitaux Universitaires de Strasbourg).

## References

1. Akiyama Y, Morioka S, Wakimoto Y, et al. Non-COVID-19 Patients with Life-threatening Diseases Who Visited a Fever Clinic: A Single-center, Observational Study in Tokyo, Japan. Intern Med Tokyo Jpn 2020; 59: 3131–3133.

2. Goulabchand R, Claret P-G, Lattuca B. What if the worst consequences of COVID-19 concerned non-COVID patients? J Infect Public Health 2020; 13: 1237–1239.

3. Abraham DA, Vijayakumar TM, Rajanandh MG. Challenges of non-COVID-19 patients with chronic illness during the pandemic. J Res Pharm Pract 2020; 9: 155–157.

4. Ojetti V, Covino M, Brigida M, et al. Non-COVID Diseases during the Pandemic: Where Have All Other Emergencies Gone? Med Kaunas Lith; 56. Epub ahead of print 1 October 2020. DOI: 10.3390/medicina56100512.

5. Arrêté du 15 mars 2020 complétant l’arrêté du 14 mars 2020 portant diverses mesures relatives à la lutte contre la propagation du virus covid-19.

6. Salje H, Tran Kiem C, Lefrancq N, et al. Estimating the burden of SARS-CoV-2 in France. Science 2020; 369: 208–211.

7. Di Domenico L, Pullano G, Sabbatini CE, et al. Impact of lockdown on COVID-19 epidemic in Île-de-France and possible exit strategies. BMC Med 2020; 18: 240.

8. Données relatives à l’épidémie de COVID-19 en France : vue d’ensemble - data.gouv.fr, /fr/datasets/donnees-relatives-a-lepidemie-de-covid-19-en-france-vue-densemble/ (accessed 23 January 2021).

9. Jessop ZM, Dobbs TD, Ali SR, et al. Personal protective equipment for surgeons during COVID-19 pandemic: systematic review of availability, usage and rationing. Br J Surg 2020; 107: 1262– 1280.

10. Bersano A, Kraemer M, Touzé E, et al. Stroke care during the COVID-19 pandemic: experience from three large European countries. Eur J Neurol 2020; 27: 1794–1800.

11. Markus HS, Brainin M. COVID-19 and stroke-A global World Stroke Organization perspective. Int J Stroke Off J Int Stroke Soc 2020; 15: 361–364.

12. Huang H-H, Yen DH-T, Kao W-F, et al. Declining Emergency Department Visits and Costs During the Severe Acute Respiratory Syndrome (SARS) Outbreak. J Formos Med Assoc 2006; 105: 31– 37.

13. Rangé G, Hakim R, Motreff P. Where have the STEMIs gone during COVID-19 lockdown? Eur Heart J Qual Care Clin Outcomes. Epub ahead of print 29 April 2020. DOI: 10.1093/ehjqcco/qcaa034.

14. Siegler JE, Heslin ME, Thau L, et al. Falling stroke rates during COVID-19 pandemic at a comprehensive stroke center. J Stroke Cerebrovasc Dis 2020; 29: 104953.

15. Mariet A-S, Giroud M, Benzenine E, et al. Hospitalizations for Stroke in France During the COVID-19 Pandemic Before, During, and After the National Lockdown. Stroke 2021; STROKEAHA120032312.

16. Estimation de la population au 1er janvier 2021 | Insee, https://www.insee.fr/fr/statistiques/1893198 (accessed 20 February 2021).

17. Heiber M, Lou WYW. Effect of the SARS outbreak on visits to a community hospital emergency department. CJEM 2006; 8: 323–328.

18. Mahmassani D, Tamim H, Makki M, et al. The impact of COVID-19 lockdown measures on ED visits in Lebanon. Am J Emerg Med. Epub ahead of print 2 December 2020. DOI: 10.1016/j.ajem.2020.11.067.

19. Dean P, Zhang Y, Frey M, et al. The impact of public health interventions on critical illness in the pediatric emergency department during the SARS-CoV-2 pandemic. J Am Coll Emerg Physicians Open. Epub ahead of print 10 August 2020. DOI: 10.1002/emp2.12220.

20. Rameez F, McCarthy P, Cheng Y, et al. Impact of a Stay-at-Home Order on Stroke Admission, Subtype, and Metrics during the COVID-19 Pandemic. Cerebrovasc Dis Extra 2020; 10: 159–165.

21. Sharma M, Lioutas V-A, Madsen T, et al. Decline in stroke alerts and hospitalisations during the COVID-19 pandemic. Stroke Vasc Neurol. Epub ahead of print 27 August 2020. DOI: 10.1136/svn-2020-000441.

22. Richter Daniel, Eyding Jens, Weber Ralph, et al. Analysis of Nationwide Stroke Patient Care in Times of COVID-19 Pandemic in Germany. Stroke 2021; 52: 716–721.

23. Nogueira RG, Qureshi MM, Abdalkader M, et al. Global Impact of COVID-19 on Stroke Care and Intravenous Thrombolysis. Neurology. Epub ahead of print 25 March 2021. DOI: 10.1212/WNL.0000000000011885.

24. Helms J, Kremer S, Merdji H, et al. Neurologic Features in Severe SARS-CoV-2 Infection. N Engl J Med 2020; 382: 2268–2270.

25. Shahjouei S, Naderi S, Li J, et al. Risk of stroke in hospitalized SARS-CoV-2 infected patients: A multinational study. EBioMedicine; 59. Epub ahead of print 1 September 2020. DOI: 10.1016/j.ebiom.2020.102939.

26. Yaghi S, Ishida K, Torres J, et al. SARS-CoV-2 and Stroke in a New York Healthcare System. Stroke 2020; 51: 2002–2011.

27. Wang D, Hu B, Hu C, et al. Clinical Characteristics of 138 Hospitalized Patients With 2019 Novel Coronavirus–Infected Pneumonia in Wuhan, China. JAMA 2020; 323: 1061–1069.

28. Merkler AE, Parikh NS, Mir S, et al. Risk of Ischemic Stroke in Patients With Coronavirus Disease 2019 (COVID-19) vs Patients With Influenza. JAMA Neurol. Epub ahead of print 2 July 2020. DOI: 10.1001/jamaneurol.2020.2730.

29. Chen R, Liang W, Jiang M, et al. Risk Factors of Fatal Outcome in Hospitalized Subjects With Coronavirus Disease 2019 From a Nationwide Analysis in China. Chest 2020; 158: 97–105.

30. Nawabi J, Morotti A, Wildgruber M, et al. Clinical and Imaging Characteristics in Patients with SARS-CoV-2 Infection and Acute Intracranial Hemorrhage. J Clin Med; 9. Epub ahead of print 6 August 2020. DOI: 10.3390/jcm9082543.

31. Cheruiyot I, Sehmi P, Ominde B, et al. Intracranial hemorrhage in coronavirus disease 2019 (COVID-19) patients. Neurol Sci 2020; 1–9.

32. Tsivgoulis G, Palaiodimou L, Katsanos AH, et al. Neurological manifestations and implications of COVID-19 pandemic. Ther Adv Neurol Disord 2020; 13: 1756286420932036.

33. Rouyer O, Pierre-Paul IN, Balde AT, et al. High Prevalence of Deep Venous Thrombosis in Non-Severe COVID-19 Patients Hospitalized for a Neurovascular Disease. Cerebrovasc Dis Extra 2020; 10: 174–180.

34. Accident vasculaire cérébral. Santé Public France, https://www.santepubliquefrance.fr/maladies-et-traumatismes/maladies-cardiovasculaires-et-accident-vasculaire-cerebral/accident-vasculaire-cerebral (accessed 18 April 2021).

